# CHIASM: A Self-Supervised Visual Field Encoder for Neuro-Ophthalmology

**DOI:** 10.64898/2026.08.23.26361135

**Authors:** T Maxwell Parker, Eric K Oermann, Scott N Grossman, Rachel C Kenney

**Author notes:** Correspondence to: T Maxwell Parker, Department of Neurology, NYU Grossman School of Medicine, New York, NY, USA. These authors contributed equally as senior authors (Scott N Grossman, Rachel C Kenney).

## Abstract

**Importance:** Artificial intelligence (AI) is under active development to support diagnosis and prognostication of glaucoma from visual fields (VF). These systems do not audit for vertical-meridian-respecting field loss patterns—known sequelae of stroke, hemorrhage, and neoplasm.

**Objective:** To develop a self-supervised encoder of automated perimetry that learns anatomically interpretable visual field structure without labels and to evaluate its capacity to identify suspected neurologic VF patterns in an independent public glaucoma dataset.

**Design, Setting, and Participants:** Diagnostic study (TRIPOD+AI). Pretraining: 23,223 unlabeled Humphrey VFs (patient-grouped training split of 28,943 fields from 3,871 University of Washington patients; UWHVF, all-comers perimetry). External evaluation: Harvard-Glaucoma Fairness dataset (Harvard-GF; 3,300 patients with paired VF and OCT from a single academic center). The encoder was never exposed to Harvard-GF during training.

**Exposures:** A 128-dimensional masked autoencoder of monocular Humphrey VF pattern-deviation data, with a supervised linear classifier on vertical-midline latent dimensions trained on per-eye expert neurological/nonneurological labels.

**Main Outcomes and Measures:** Primary: classifier accuracy under hard-negative evaluation (cross-validated balanced accuracy and AUC). Secondary: held-out specificity on structurally separated UWHVF controls; expert confirmation and OCT structural correlates of classifier-identified Harvard-GF suspects.

**Results:** Masked reconstruction recovered structure concordant with retinal neuroanatomy; 50 of 128 latent dimensions emerged spatially specialized (vs 23 for the TD encoder). Under hard-negative evaluation the classifier achieved cross-validated balanced accuracy 0.78 (95% CI, 0.75-0.82) and AUC 0.85 (95% CI, 0.82-0.89), with no false positives among 100 held-out controls. Applied to Harvard-GF without fine-tuning, it identified a top-20 of 1,748 glaucoma- labeled patients (1.1%) with morphology inconsistent with glaucoma; all 20 were NHT-positive (mean 62.4), and OCT showed preserved superior (Cohen d = +0.68; P < .001) and inferior (d = +0.63; P = .003) RNFL versus severity-matched controls.

**Conclusions and Relevance:** A self-supervised VF encoder learned anatomically interpretable visual field structure from unlabeled data and identified suspected neurological cases in a curated glaucoma dataset, with expert, rule-based, and OCT corroboration. Visual field datasets used to train glaucoma AI may benefit from neurological screening; the encoder reported here supports such audits and provides a foundation for neuro-ophthalmic AI beyond fundus photography and OCT.

**Key Points:** *Question:* Can self-supervised learning learn anatomically interpretable visual field structure from unlabeled visual fields and identify neurological field loss?

*Findings:* Trained on 28,943 unlabeled visual fields, the encoder learned anatomically interpretable visual field structure and revealed ≥20 of 1,748 patients (≥1.1%) in an external glaucoma dataset with vertical-midline loss and retinal nerve fiber layer preservation.

*Meaning:* A self-supervised encoder learned anatomically interpretable visual field structure, and can audit glaucoma datasets for neurological field loss, reducing the risk that future AI misclassifies intracranial disease.

## Introduction

Automated perimetry detects functional correlates of pathology across the entire visual pathway — optic neuropathy, chiasmal compression, retrochiasmal disease from stroke, hemorrhage, or tumor — and is performed daily in eye and neurology clinics worldwide. Deep learning has transformed other domains of ophthalmic imaging: dozens of artificial intelligence (AI) systems interpret glaucomatous visual fields,^1–6^ foundation models trained on fundus photographs generalize across retinal disease,^7^ and the US FDA has approved automated fundus interpretation for diabetic retinopathy screening.^8^ No modern correlate exists for neuro-ophthalmic visual field (VF) interpretation: computational work remains limited to rule-based and shallow-learning approaches,^9^ and no learned representation of VF morphology has been published.

The gap is clinical, not merely computational. A significant minority (6-23%) of patients carrying a diagnosis of normal-tension glaucoma harbor intracranial lesions on neuroimaging,^10,11^ with up to eight-year delays before neurologic diagnosis.^12^ A quarter of non-glaucomatous optic neuropathies resembling glaucoma on fundoscopy are misdiagnosed, prolonging unnecessary pressure-lowering treatment.^13,14^ Field loss from stroke, hemorrhage, or tumor — or bitemporal loss from a pituitary adenoma compressing the chiasm — is morphologically distinct from glaucomatous neuropathy, yet the automated outputs used clinically (mean deviation, glaucoma hemifield test, pattern standard deviation) grade severity attributable to glaucoma rather than differentiating etiologies. The shortage of neuro-ophthalmic providers compounds the problem.^15,16^

Existing approaches capture one facet of the issue. The Boland Neurological Hemifield Test (NHT) scores vertical-midline asymmetry through rule-based pattern-deviation thresholds and achieves an area under the curve of 0.90,^17^ extended by McCoy and colleagues to incorporate binocular concordance.^18^ A shallow neural network distinguishes chiasmal from glaucomatous field loss at sensitivity and specificity above 95%,^19^ and archetypal analysis identifies hemianopic patterns within glaucoma repositories.^20^ None produces a continuous embedding (a continuous numerical representation) that operates without labels, generalizes across datasets, supports phenotype-specific probing, or aligns cross-modally with structural imaging.

The absence of neuro-ophthalmic VF AI shapes how glaucoma AI is built. The American Academy of Ophthalmology defines glaucoma diagnosis as requiring an anatomically open angle, characteristic optic nerve cupping, retinal nerve fiber layer thinning, typical VF defects, and exclusion of secondary causes including neurologic disease.^21^ VF criteria alone do not meet this standard, yet public AI training datasets are routinely labeled by VF criteria alone^1,22^ and downstream studies use these labels as ground truth.^1–3^ When neurological patients enter such corpora, glaucoma models can learn their field loss — the morphological signature of stroke or intracranial mass — as glaucomatous.

We present CHIASM (Cross-Hemifield Identification by Autonomous Self-supervised Masking), a masked autoencoder trained on unlabeled visual fields that represents each monocular Humphrey VF as 128 numbers (an embedding). We characterize what the encoder learns, evaluate a supervised linear classifier over its latent dimensions, and apply that classifier to an independent curated glaucoma dataset to test whether the representation surfaces neurological field loss outside its training institution.

## Methods

This study is reported in accordance with the TRIPOD+AI statement.^23^

### Datasets

UWHVF (University of Washington Humphrey Visual Fields) comprises 28,943 Humphrey VFs from 3,871 patients — all-comers perimetry with no diagnostics labels.^24^ UWHVF is not a glaucoma dataset; it contains patients with all etiologies of visual field loss and provides bilateral pairs for 3,557 patients.

Harvard-GF (Harvard-Glaucoma Fairness) comprises 3,300 patients with paired VF and optical coherence tomography (OCT) retinal nerve fiber layer (RNFL) data from a single academic center.^22^ Glaucoma labels were assigned by Wang and colleagues using VF criteria alone: glaucoma as mean deviation (MD) worse than −3 dB with abnormal glaucoma hemifield test and pattern standard deviation; non-glaucoma as MD at or better than −1 dB with both normal. The cohort contains 1,748 glaucoma-labeled and 1,552 non-glaucoma-labeled patients (one eye per patient, no laterality stored). UWHVF fields are stored in a canonical orientation, laterality being identifiable from the physiologic blind spot; Harvard-GF distributes none. Because the classifier operates on a signed nasal-temporal axis, we verified orientation empirically: the mean nasal-minus-temporal total-deviation profile matched UWHVF’s (r = 0.97), consistent with a uniformly oriented cohort.

To build an evaluation set enriched for neurological patterns, we screened UWHVF with the Boland NHT^17^ and identified 292 patients with NHT ≥ 30 on at least one field. All 2,640 fields from these patients were reviewed eye-by-eye by the first author (T.M.P., neurologist) with TD and PD side-by-side and each eye labeled neuro_suspect, not_neuro_suspect, or uncertain, yielding 889 neuro_suspect fields (221 eyes, 130 patients) and 966 not_neuro_suspect fields (184 eyes, 114 patients). Because both classes came from NHT-flagged patients, the negative class comprises eyes that triggered rule-based review but lacked neurological morphology on inspection — a hard-negative design. Per-fold composition is in eMethods and eTable 4. This study analyzed publicly available, de-identified datasets and did not constitute human subjects research requiring institutional review board approval.

Monocular masked autoencoder:

We developed a self-supervised model of monocular Humphrey visual fields that converts each field into 128 features learned from unlabeled data. Development proceeded in three stages with disjoint data (Figure 1): Stage 1, self-supervised pretraining on 23,223 unlabeled UWHVF fields; Stage 2, a supervised linear classifier fit on eye-level expert labels under 5-fold patient-grouped cross-validation (all fields from a patient in one fold); Stage 3, external application to Harvard-GF, which contributed no data at any prior stage.

**Figure 1:**
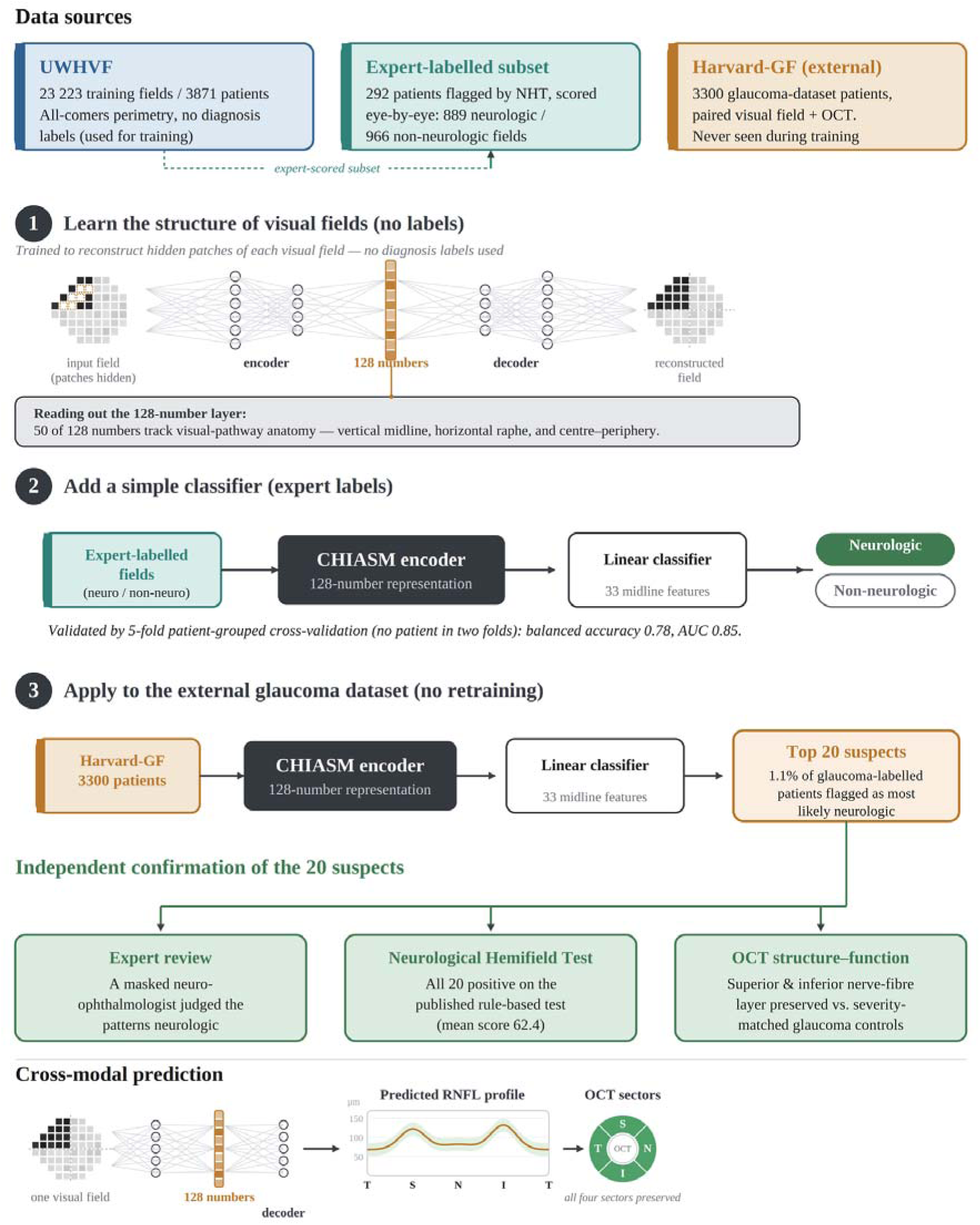
Study workflow. Schematic of the four-stage CHIASM pipeline with color-coded datasets and disjoint data at every stage. Stage 1 (self-supervised learning, no labels): a masked autoencoder is trained on the 23,223-field training split of the University of Washington Humphrey Visual Fields (UWHVF); portions of each field are hidden and reconstructed, and the encoder compresses each field into a 128-number representation (embedding), 50 of 128 dimensions of which align with visual-pathway anatomy. Stage 2 (supervised classifier, expert labels): a linear classifier on 33 vertical-midline dimensions of the frozen encoder is trained on expert-labeled neurologic versus non-neurologic eye-fields and validated by 5-fold patient-grouped cross-validation (balanced accuracy 0.78, AUC 0.85). Stage 3 (external application, no retraining): the frozen encoder and classifier are applied to 3,300 Harvard-Glaucoma Fairness (Harvard-GF) patients, flagging the 20 highest-scoring (1.1%) as most suggestive of a neurological pattern. Independent confirmation: the 20 suspects are corroborated by masked expert review by a neuro-ophthalmologist, the Boland Neurological Hemifield Test (all 20 positive; mean 62.4), and OCT structure-function analysis (preserved superior and inferior retinal nerve fiber layer versus severity-matched glaucoma controls). A cross-modal decoder additionally predicts the peripapillary RNFL profile from the visual field embedding alone; in the held-out suspects, predicted RNFL was preserved in all four sectors.

Pattern deviation (PD) was computed deterministically from total deviation (TD) using the Humphrey Field Analyzer 7th-highest rule. We trained six encoders in a 2×3 factorial design crossing input representation (TD, PD, TD+PD) with positional-encoding initialization (retinal- coordinate, random), plus a no-positional control. Each of the 128 latent dimensions was categorized by differential activation between anatomically defined hemifield regions (vertical- midline, horizontal-midline, central-peripheral, severity, or mixed). The PD coordinate- initialized encoder was selected as primary: 50 of 128 dimensions were spatially specialized versus 23 for the TD encoder.

### Model architecture and training

The encoder is a 4-layer transformer (model dimension 128, 4 attention heads, pre-norm, dropout 0.1) that treats each of the 52 test locations as one token and mean-pools the final layer into a 128-number representation (537,088 parameters). Positional encodings were initialized to retinal coordinates; random-initialization and no-positional variants were trained for comparison. In pretraining, a random 30% to 50% of locations was hidden and a 2-layer decoder reconstructed the hidden values under a Huber loss^25^; the decoder was then discarded. Optimization used AdamW with cosine decay and early stopping on validation reconstruction loss, with a single random seed; checkpoints are not bit-reproducible. Training used the 23,223 UWHVF training-split fields, with reconstruction evaluated on 2,854 held-out fields. Full specification is in eMethods.

### Neurological proximity score

To identify visual fields suspicious for neurologic loss, we trained a supervised classifier (logistic regression) on the 33 vertical-midline latent dimensions of the primary encoder, using the expert-labeled eye-fields. These dimensions were chosen for interpretability: each corresponds to a clinically recognizable vertical-midline-respecting pattern with anatomically inspectable weights. Performance was evaluated by 5-fold patient-grouped cross-validation under hard-negative (NHT > 30) evaluation. Specificity was assessed on 100 held-out UWHVF patients disjoint from training (50 clean-normal, 50 clean-glaucoma); at the threshold later used to flag Harvard-GF suspects, none were flagged. To test whether performance depended on the vertical-midline dimensions rather than feature count or raw-field access, we compared the classifier against the complementary 95-dimension subspace, random 33-dimension subsets, raw 52-point PD values and their principal-component decomposition with a linear model, and the NHT score alone. The vertical-midline subspace was selected for interpretability rather than unique sufficiency, and the primary encoder on anatomical specialization rather than accuracy.

### Cross-dataset screening

All 3,300 Harvard-GF patients were encoded by the primary UWHVF-trained encoder (no fine-tuning) and scored by the vertical-midline classifier, with zero Harvard-GF exposure at any stage. To form a conservative review cohort, we set the operating threshold at the score of the 20th-highest-ranked patient. Expert review was performed by a board-certified neuro-ophthalmologist (S.N.G.) with TD and PD heatmaps side-by-side, judging whether cases warranted further neurological investigation regardless of coexisting glaucoma.

### NHT implementation

We implemented the Boland NHT^17^ and McCoy binocular extension.^18^ Because the NHT’s per-location probability thresholds derive from a proprietary normative reference, we estimated them empirically from 4,532 UWHVF normals (MD > −2 dB) at each of 52 locations (probability levels <5%, <2%, <1%, <0.5%). The published cutoff, an NHT score of 30, was used throughout.^17^

### OCT structure-function analysis

Sectoral RNFL thickness from Harvard-GF OCT was compared between the top-20 suspects and severity-matched glaucoma controls, sectors following the Garway-Heath convention^26^ (±45° arcs around the cardinal peripapillary positions). Controls were glaucoma-labeled patients unlikely to carry a neurological pattern (classifier rank below the top 200, NHT score < 28), matched 5:1 on mean deviation (±2 dB) so comparisons reflect etiology rather than severity. RNFL residual was actual minus that predicted by linear regression of sectoral RNFL on MD across the full Harvard-GF glaucoma cohort.

### Statistical Methods

Classification performance was estimated by 5-fold stratified cross-validation grouped by patient, so all fields from a patient fell within a single fold. Balanced accuracy and area under the receiver operating characteristic curve were computed on pooled out-of-fold predictions, with patient-level bootstrap CIs (5,000 iterations, percentile method). Across-fold standard deviations are reported as between-fold dispersion, not confidence intervals. Held-out specificity is reported with exact binomial bounds. Sectoral RNFL differences were assessed by Mann-Whitney U with Cohen d and 3,000-iteration bootstrap CIs, against an empirical null from severity-matched glaucoma subsets. Correlations are Spearman coefficients. Analyses used Python (PyTorch, scikit-learn, SciPy). Two-sided P < .05 was considered significant; P values are descriptive.

### Findings

#### Self-supervised reconstruction recovers structure concordant with retinal neuroanatomy

Masked reconstruction alone was sufficient to recover structure concordant with the major axes of retinal neuroanatomy (Figure 2; eTable 1, eTable 3, eFigure 2). The primary encoder — pattern-deviation input, retinal coordinate–initialized positional encodings — developed 50 of 128 latent dimensions as spatially specialized. Dimensions could satisfy more than one axis, yielding 70 axis assignments across these 50: 33 vertical-midline, 18 horizontal-raphe, and 19 central-peripheral. The equivalent total-deviation encoder developed only 23. Pattern-deviation preprocessing — which removes the generalized depression component and concentrates signal on focal pattern — drove this 2.2-fold gain. A no-positional control collapsed to severity-only features (zero central-peripheral dimensions; reconstruction MAE 3.12 dB versus 1.96 dB), confirming positional information was necessary.

**Figure 2:**
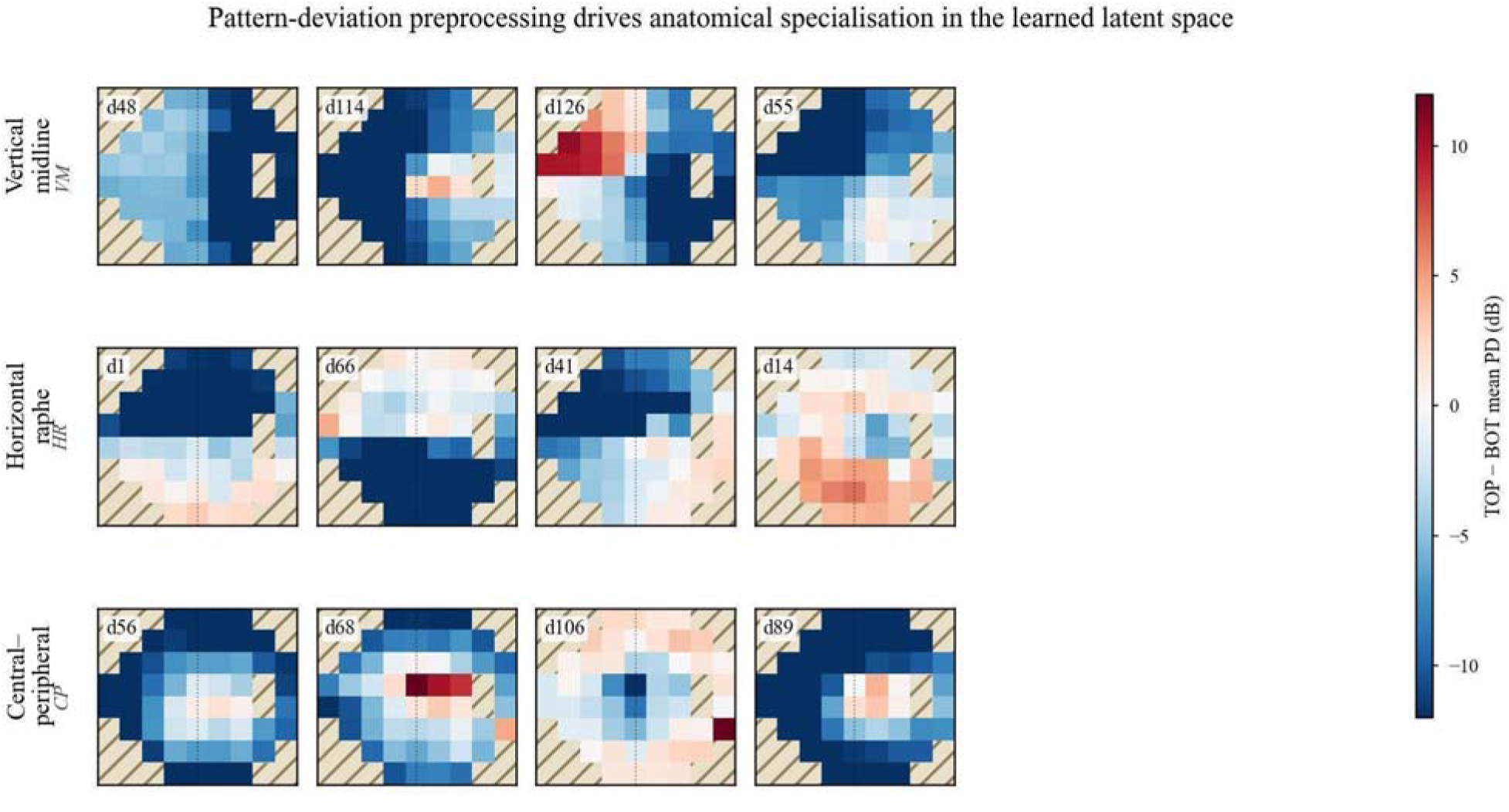
Pattern-deviation preprocessing drives anatomical specialization in the learned latent space. Per-dimension activation contrast maps from the primary encoder (pattern- deviation input, retinal coordinate-initialised positional encoding). Procedure: 5,000 UWHVF training fields were encoded; for each dimension (one of the 128 scalar outputs of the model), the 50 fields most strongly activating and the 50 most strongly suppressing it were identified, and their pattern-deviation maps averaged. Each panel shows the per-location difference between top-50 and bottom-50 mean PD (dB) — a 52-point spatial contrast in clinician-readable input units. Blue indicates locations darker (more defect-like) when the dimension fires high; red indicates locations relatively preserved. The same per-dimension diff map, reduced to a between-hemifield mean difference, defines the categorization criterion in Methods (≥ 5 dB on a spatial axis). Top row: vertical-midline-specialized dimensions (4 of 33). Middle row: horizontal-raphe-specialized dimensions (4 of 18). Bottom row: central-peripheral-specialized dimensions (4 of 19). No anatomical or diagnostic supervision was provided during pretraining.

Beyond the three primary anatomical axes, the encoder’s latent space supports hand-specified classifiers for phenotypes spanning the visual pathway from optic nerve to occipital cortex (Figure 3). Each is an unweighted sum over a curated set of latent dimensions, specified from anatomical reasoning rather than fitted to labels, and presented as a demonstration of the latent space’s queryability rather than as a validated classifier. Ten canonical phenotypes — centrocecal scotoma, peripheral constriction, bitemporal hemianopia, left and right homonymous hemianopia, left and right Meyer’s-loop and superior-radiation quadrantanopias, and bilateral central scotoma — each occupy confined regions of the encoder’s UMAP (uniform manifold approximation and projection) projection over a 5,000-eye UWHVF background, indicating pathway-localized morphology is locally structured in the representation.

**Figure 3:**
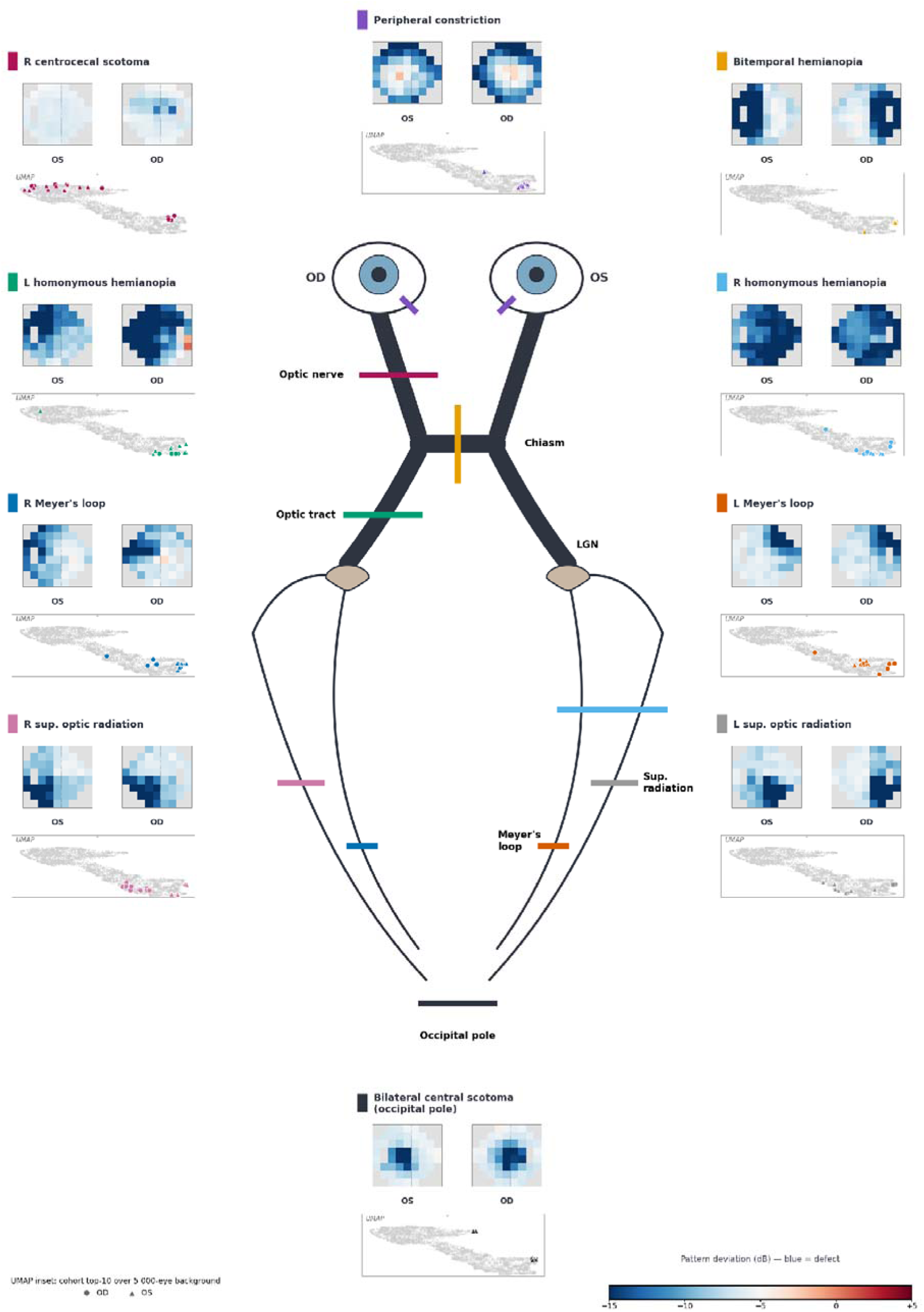
Pathway-localised phenotype classifier library. The encoder’s latent space supports phenotype-specific classifiers spanning the visual pathway from optic nerve to occipital cortex. Central schematic: visual pathway from globe to occipital pole, with color-coded annotations marking the anatomical origin of each phenotype (chiasm, lateral geniculate, Meyer’s loop, superior optic radiation, occipital pole). Peripheral panels: ten canonical neuro- ophthalmic VF phenotypes, each shown with paired OS and OD pattern-deviation thumbnails of the top-10 cohort mean (pattern-deviation dB scale at bottom right; blue indicates defect). Pathway locations are clinical annotations of each phenotype’s expected lesion site. UMAP insets: per-phenotype top-10 cohort exemplars (circles: OD; triangles: OS) projected onto a 2D UMAP embedding computed from the encoder’s 128-dimensional latent representation over a 5,000-eye UWHVF background (grey). Each phenotype’s exemplars localise to a confined region of the UMAP, demonstrating that pathway-specific VF morphology occupies distinct neighbourhoods of the encoder’s representation and that the encoder supports phenotype-specific probing without retraining.

#### Cross-institutional screening identifies neurological suspects

The supervised vertical-midline classifier was applied to the external Harvard-GF dataset without fine-tuning, identifying a top-20 cohort of glaucoma-labeled patients (1.1%; Figure 4; ranks 21-50 in eFigure 4) whose VF morphology was less consistent with glaucomatous etiology and would, in modern practice, prompt consideration of further workup such as with neuroimaging. Under hard-negative (NHT > 30) evaluation, the classifier achieved balanced accuracy 0.78 (95% CI, 0.75-0.82; sensitivity 0.70, specificity 0.86) and AUC 0.85 (95% CI, 0.82- 0.89) (Table 1; eFigure 3), with perfect specificity on 100 held-out UWHVF controls. Simple alternatives on the same folds performed less well: principal-component decomposition of the raw PD values with a linear model (0.69; AUC 0.73), logistic regression on the raw values (0.69; AUC 0.74), and the NHT score alone (0.67; AUC 0.74). Blind expert review by a board-certified neuro-ophthalmologist (S.N.G.) judged the patterns suspicious for neurological field loss and warranting investigation. Top-20 patients had mean NHT 62.4 (range 32–80) and mean MD −12.9 dB (range −21.0 to −5.5). Classifier score correlated weakly and positively with MD within the glaucoma subset (Spearman ρ = +0.17; P < .001) — higher-scoring patients had slightly milder loss, so the ranking does not reduce to severity.

**Figure 4:**
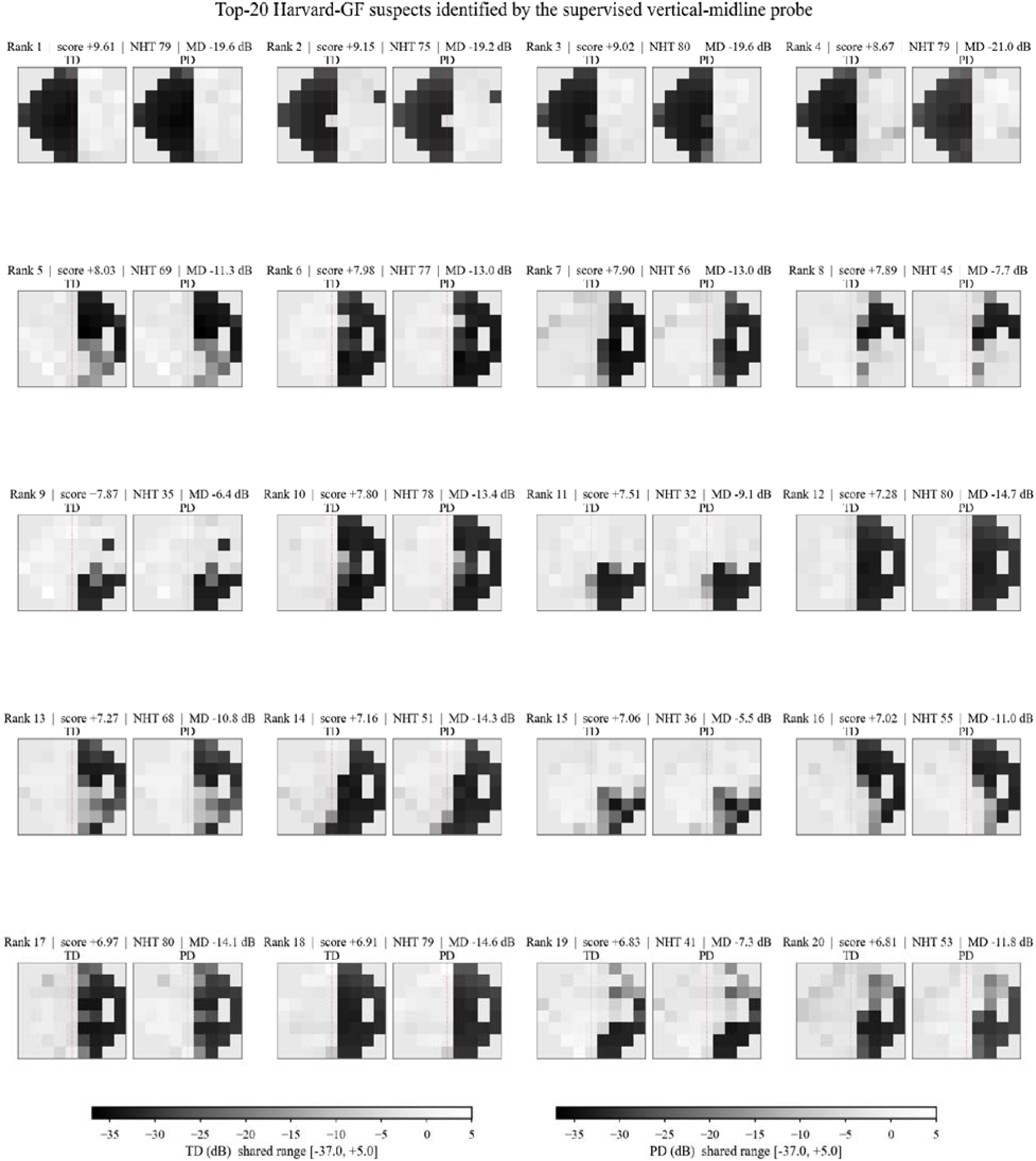
Top-20 Harvard-GF suspects identified by the supervised vertical-midline classifier. Visual field heatmaps of the 20 glaucoma-labeled Harvard-GF patients ranked highest by the supervised vertical-midline classifier. Each patient is shown with paired total-deviation (left) and pattern-deviation (right) greyscale plots, annotated with classifier score (signed log- odds), NHT score at the published cutoff of 30, and mean deviation. All 20 show vertical- midline-respecting hemifield morphology; blinded expert review by a board-certified neuro-ophthalmologist (SNG) confirmed suspected neurological patterns less consistent with glaucomatous optic neuropathy.

**Table 1:** Supervised vertical-midline classifier validation and feature-subspace ablation Panel A reports validation of the supervised vertical-midline classifier (logistic regression on the 33 vertical- midline-categorized dimensions of the primary encoder, trained to separate expert-labeled neuro_suspect from not_neuro_suspect eye-fields). Cross-validation uses 5-fold stratified group splits with grouping by patient_id, under hard-negative (NHT > 30) evaluation; held-out specificity controls are 50 clean-normal and 50 clean- glaucoma patients disjoint from the training pool.

| Panel A — Classifier validation | Value |
| --- | --- |
| Balanced accuracy, pooled out-of-fold | 0.78 (95% CI, 0.75-0.82) |
| AUC, pooled out-of-fold | 0.85 (95% CI, 0.82-0.89) |
| Sensitivity, pooled out-of-fold | 0.70 (95% CI, 0.64-0.77) |
| Specificity, pooled out-of-fold | 0.86 (95% CI, 0.83-0.90) |
| Per-fold balanced accuracy (between-fold dispersion) | 0.70, 0.77, 0.83, 0.85, 0.77 |
| Held-out clean-normal specificity (n = 50; NHT < 10, MD > -3 dB) | 0 of 50 flagged at top-20 threshold (95% CI for false-positive rate, 0-5.8%); max score -0.77 |
| Held-out clean-glaucoma specificity (n = 50; NHT < 10, MD < -5 dB, low hemianopic loading) | 0 of 50 flagged at top-20 threshold (95% CI for false-positive rate, 0-5.8%) |
| Top-20 operating threshold (signed log-odds) | ≥ 6.807 |
| Harvard-GF top-20 cohort composition | all glaucoma-labeled; mean NHT 62.4 (range 32-80); mean MD -12.9 dB (range -21.0 to -5.5) |

#### NHT independently validates the classifier’s top-20

The Boland NHT at the published cutoff of 30 independently flagged all 20 classifier-identified suspects, with mean NHT 62.4. The two methods converge on the same patients through structurally different mechanisms — a continuous signed log-odds in a learned representation, and a rule-based asymmetry statistic from pattern-deviation probabilities — corroborating the top-20 without reliance on a single modality.

#### OCT structural preservation supports non-glaucomatous etiology

Top-20 suspects showed preserved retinal nerve fiber layer thickness relative to severity-matched glaucoma controls in the superior (Δ = +20.1 µm; Cohen d = +0.68; P < .001) and inferior (Δ = +17.2 µm; d = +0.63; P = .003) sectors, with temporal and nasal sectors null (eTable 2, eFigure 1). Both effects exceeded the 95% CI of a bootstrap null from 100 random severity-matched subsets. This pattern — severe field loss with preserved superior and inferior RNFL and no temporal or nasal difference from matched controls — is a structure-function dissociation classically seen in retrochiasmal disease.

#### Cross-modal VF-to-OCT reconstruction

A cross-modal decoder predicted the 256-point peripapillary RNFL profile (TSNIT) from VF embeddings, using Harvard-GF patients outside the top-200 rank (patient-level splits; top-20 excluded). Across 24 encoder × decoder combinations, global mean absolute error clustered at 23.3–23.7 µm — a representation-invariant ceiling set by 52 VF locations against 256 OCT positions under test-retest variability. Applied to the held-out top-20, predicted RNFL was preserved in every sector (eFigure 5): temporal d = +1.59, superior +1.40, nasal +1.78, inferior +1.37, global +1.69, all exceeding the null 95% CI. Predicted preservation extends to sectors null on actual OCT, indicating a morphology-level signal rather than sector-specific concordance.

## Discussion

CHIASM is a self-supervised model for neuro-ophthalmic visual field analysis: The encoder learned anatomically interpretable structure from unlabeled data, distinguished neurological from non-neurological field loss with a simple supervised classifier, and linked VF to OCT-derived RNFL.

At a conservative operating threshold, the classifier identified the top 20 glaucoma-labeled cases (1.1%) as most suggestive of a neurological pattern; all were NHT-positive with OCT structure-function dissociation. These findings do not estimate contamination prevalence; the threshold was chosen for clarity. The implication nonetheless extends beyond this dataset. The American Academy of Ophthalmology defines glaucoma diagnosis as requiring characteristic optic nerve and RNFL findings together with exclusion of secondary causes, including neurological disease.^21^ Visual-field criteria alone cannot satisfy that definition: they describe the pattern of loss, not its cause. Public AI corpora are nonetheless routinely labeled by VF criteria alone, and downstream studies treat those labels as ground truth.^1–3^ Every model trained on such a corpus inherits a label that is not the diagnosis it purports to represent: neurological field loss is learned as glaucomatous, and the misclassification is invisible to internal validation because the test labels share the same definition. This is a field-level problem rather than a property of any single dataset, and it is tractable: screening of the kind demonstrated here can be applied before training, and etiologic confirmation required of new corpora.

The same blind spot extends to care delivery. Technician-delivered virtual glaucoma clinics operate across the UK National Health Service, collecting visual field and OCT data for asynchronous specialist review,^27–30^ and are a natural entry point for AI-assisted triage. Because those pathways triage on the same etiology-agnostic VF measures used to label research datasets, a screening step for neurological morphology belongs upstream of any automated classification.

Masked reconstruction hides portions of each VF and learns to predict them (Figure 2), which requires representing spatial structure. From this objective alone — without anatomical labels — the encoder allocated 50 of 128 dimensions to vertical-midline, horizontal-raphe, and central-peripheral structure, versus 23 for the TD encoder. That a representation trained only on UWHVF transferred zero-shot to Harvard-GF — a different institution, population, and era — indicates structure that generalizes rather than dataset-specific artifact.

The NHT and the classifier implement the same detection task — flagging vertical-midline asymmetry — as a hand-engineered rule and a learned discriminator on 33 interpretable dimensions; both converge on the maxim that neuro-ophthalmologists “worship the vertical meridian.” The NHT serves here as corroboration rather than competition.

The encoder’s 128 dimensions are a general-purpose substrate for neuro-ophthalmic VF morphology. The vertical-midline classifier is one instance; other subspaces address altitudinal defects, central scotomas, quadrantanopias and complex paracentral “cloverleaf” patterns (Figure 3), and potentially functional (non-organic) fields, paralleling supervised detection of functional seizures from records.^31^ Any such classifier fits on a small labeled set atop the frozen encoder, and the same embedding decodes to predicted RNFL reproducing the dissociation in held-out suspects. Archetypal analysis,^20^ the closest prior method, decomposes each VF onto fixed templates and supports none of these capabilities.

OCT adds an independent structural modality: top-20 suspects showed RNFL preservation classically seen in retrochiasmal disease not in glaucoma, and a decoder trained without these patients predicted preserved RNFL across all sectors (global Cohen d = +1.69). Interpretation is bounded by inferred (not clinically confirmed) etiology and unknown disease duration, which governs retrograde RNFL thinning over months to years.^32,33^

### Limitations

Harvard-GF suspects were not validated against clinical records; retrospective validation of similar data at our institution is in progress. Only two US academic institutions contribute open public VF data, limiting external validation. Harvard-GF distributes no laterality field, so per-case assignment of defects to anatomical nasal or temporal hemifields could not be verified; the classifier is orientation-sensitive, and the cohort-level check supports but does not document consistent orientation. Monocular input precludes binocular-phenotype confirmation: a monocular nasal-hemifield defect is not reliably distinguished from a nasal quadrantanopia without fellow-eye data, so model-identified cases indicate patterns warranting clinical review rather than confirmed diagnoses. The VF-to-OCT prediction ceiling near 23 µm precludes per-patient fidelity; predicted-RNFL dissociation reached significance at the group level only.

## Conclusions

CHIASM establishes that a self-supervised masked autoencoder recovers clinically meaningful structure from unlabeled visual fields and supports interpretable classification, dataset auditing for neurologic field loss, and cross-modal structural prediction. Future work will pursue prospective validation, larger and more diverse datasets, clinically confirmed cases, and pilot integration of screening into automated care pathways.

## Supporting information

TRIPOD AI Checklist

## Data Availability

All data produced are available online at https://github.com/uw-biomedical-ml/uwhvf (UWHVF) and https://github.com/Harvard-Ophthalmology-AI-Lab/Harvard-GF (Harvard-GF)

https://github.com/uw-biomedical-ml/uwhvf

## Acknowledgments

None reported.

## Author Contributions

Dr Parker had full access to all of the data in the study and takes responsibility for the integrity of the data and the accuracy of the data analysis. All authors approved the final manuscript and accept accountability for the work.

Concept and design: Parker, Grossman, Kenney.

Acquisition, analysis, or interpretation of data: Parker, Grossman, Kenney, Oermann.

Drafting of the manuscript: Parker, Grossman, Kenney.

Critical review of the manuscript for important intellectual content: All authors.

Statistical analysis: Parker, Kenney.

Administrative, technical, or material support: Grossman, Kenney

Supervision: Grossman, Kenney.

## Data Sharing Statement

UWHVF is publicly available under the BSD 3-Clause license. Harvard-GF is publicly available under the CC BY-NC-ND 4.0 license. Model weights, code, and the expert-labeled evaluation set are available from the corresponding author upon reasonable request.

## References

1. Shi M, Luo Y, Tian Y, et al. Equitable artificial intelligence for glaucoma screening with fair identity normalization. npj Digit Med. 2025;8:46.

2. Afolabi SO, Gheisi L, Shan J, Shen LQ, Wang M, Shi M. Equity-enhanced glaucoma progression prediction from OCT with knowledge distillation. npj Digit Med. 2025;8:477.

3. Luo Y, Tian Y, Shi M, et al. FairCLIP: harnessing fairness in vision-language learning. In: Proceedings of the IEEE/CVF Conference on Computer Vision and Pattern Recognition 2024: 12289–301.

4. Wang M, Shen LQ, Pasquale LR, et al. An artificial intelligence approach to detect visual field progression in glaucoma based on spatial pattern analyzis. Invest Ophthalmol Vis Sci. 2019;60:365–75.

5. Wu S, Chen JY, Mohammadzadeh V, et al. Self-supervised denoising of visual field data improves detection of glaucoma progression. Preprint 2024. https://arxiv.org/abs/2411.12146.

6. Berchuck SI, Mukherjee S, Medeiros FA. Estimating rates of progression and predicting future visual fields in glaucoma using a deep variational autoencoder. Sci Rep. 2019;9:18113.

7. Zhou Y, Chia MA, Wagner SK, et al. A foundation model for generalizable disease detection from retinal images. Nature. 2023;622:156–63.

8. Teng CW, Patel SD, Barkmeier AJ, et al. Autonomous artificial intelligence in diabetic retinopathy testing—lessons learned on successful health system adoption. Ophthalmol Sci. 2025;5(5):100789.

9. Kenney RC, Requarth TW, Jack AI, Hyman SW, Galetta SL, Grossman SN. AI in neuro-ophthalmology: current practice and future opportunities. J Neuroophthalmol. 2024;44:308–18.

10. Ahmed IIK, Feldman F, Kucharczyk W, Trope GE. Neuroradiologic screening in normal-pressure glaucoma: study results and literature review. J Glaucoma. 2002;11:279–86.

11. Kosior-Jarecka E, Wróbel-Dudzińska D, Pietura R, et al. Results of neuroimaging in patients with atypical normal-tension glaucoma. BioMed Res Int. 2020;2020:9093206.

12. Drummond SR, Weir C. Chiasmal compression misdiagnosed as normal-tension glaucoma: can we avoid the pitfalls? Int Ophthalmol. 2010;30:215–19.

13. Dias DT, Ushida M, Battistella R, Dorairaj S, Prata TS. Neurophthalmological conditions mimicking glaucomatous optic neuropathy: analyzis of the most common causes of misdiagnosis. BMC Ophthalmol. 2017;17:2.

14. Choudhari NS, Neog A, Fudnawala V, George R. Cupped disc with normal intraocular pressure: the long road to avoid misdiagnosis. Indian J Ophthalmol. 2011;59:491–97.

15. DeBusk A, Subramanian PS, Scannell Bryan M, Moster ML, Calvert PC, Frohman LP. Mismatch in supply and demand for neuro-ophthalmic care. J Neuroophthalmol. 2022;42:62–67.

16. Mollan SP, Menon V, Cunningham A, et al. Neuro-ophthalmology in the United Kingdom: providing a sustainable, safe and high-quality service for the future. Eye. 2024;38:2235– 37.

17. Boland MV, McCoy AN, Quigley HA, et al. Evaluation of an algorithm for detecting visual field defects due to chiasmal and postchiasmal lesions: the neurological hemifield test. Invest Ophthalmol Vis Sci. 2011;52:7959–65.

18. McCoy AN, Quigley HA, Wang J, et al. Development and validation of an improved neurological hemifield test to identify chiasmal and postchiasmal lesions by automated perimetry. Invest Ophthalmol Vis Sci. 2014;55:1017–23.

19. Thomas PBM, Chan T, Nixon T, Muthusamy B, White A. Feasibility of simple machine learning approaches to support detection of non-glaucomatous visual fields in future automated glaucoma clinics. Eye. 2019;33:1133–39.

20. Elze T, Pasquale LR, Shen LQ, Chen TC, Wiggs JL, Bex PJ. Patterns of functional vision loss in glaucoma determined with archetypal analyzis. J R Soc Interface. 2015;12(103):20141118.

21. Gedde SJ, Vinod K, Wright MM, et al. Primary Open-Angle Glaucoma Preferred Practice Pattern. Ophthalmology. 2021;128:P71–P150.

22. Luo Y, Tian Y, Shi M, et al. Harvard Glaucoma Fairness: a retinal nerve disease dataset for fairness learning and fair identity normalization. IEEE Trans Med Imaging. 2024;43:2623–33.

23. Collins GS, Moons KGM, Dhiman P, et al. TRIPOD+AI statement: updated guidance for reporting clinical prediction models that use regression or machine learning methods. BMJ. 2024;385:e078378.

24. Montesano G, Chen A, Lu R, Lee CS, Lee AY. UWHVF: a real-world, open source dataset of perimetry tests from the Humphrey Field Analyzer at the University of Washington. Transl Vis Sci Technol. 2022;11:2.

25. He K, Chen X, Xie S, Li Y, Dollár P, Girshick R. Masked autoencoders are scalable vision learners. In: Proceedings of the IEEE/CVF Conference on Computer Vision and Pattern Recognition 2022: 16000–09.

26. Garway-Heath DF, Poinoosawmy D, Fitzke FW, Hitchings RA. Mapping the visual field to the optic disc in normal tension glaucoma eyes. Ophthalmology. 2000;107(10):1809–1815.

27. Kotecha A, Baldwin A, Brookes J, Foster PJ. Experiences with developing and implementing a virtual clinic for glaucoma care in an NHS setting. Clin Ophthalmol. 2015;9:1915–1923.

28. Kotecha A, Bonstein K, Cable R, Cammack J, Clipston J, Foster P. Qualitative investigation of patients’ experience of a glaucoma virtual clinic in a specialist ophthalmic hospital in London, UK. BMJ Open. 2015;5:e009463.

29. Jayaram H, Strouthidis NG, Gazzard G. The COVID-19 pandemic will redefine the future delivery of glaucoma care. Eye. 2020;34:1203–1205.

30. Kotecha A, Brookes J, Foster PJ. A technician-delivered virtual clinic for triaging low-risk glaucoma referrals. Eye. 2017;31:899–905.

31. Kerr WT, McFarlane KN, Pucci GF, et al. Supervised machine learning compared to large language models for identifying functional seizures from medical records32. Epilepsia. 2025;66(4):1163–1174.

32. Jindahra P, Petrie A, Plant GT. Retrograde trans-synaptic retinal ganglion cell loss identified by optical coherence tomography. Brain. 2009;132:628–34.

33. Mitchell JR, Oliveira C, Tsiouris AJ, Dinkin MJ. Corresponding ganglion cell atrophy in patients with postgeniculate homonymous visual field loss. J Neuroophthalmol. 2015;35:353–59.

