## Supplemental Figures for "CHIASM: A Self-Supervised Visual Field Encoder for Neuro-Ophthalmology"

### eMethods

**Dimension activation analysis.** Each of 128 latent dimensions in each of the six encoders was tested for differential activation between anatomically defined hemifield regions: nasal versus temporal (vertical midline), superior versus inferior (horizontal raphe), and central versus peripheral (central-peripheral gradient). For each dimension, mean absolute activation was computed separately within each region over the UWHVF training set. A dimension was categorized as specialized on a given axis when the between-region activation difference exceeded 5 dB; a single dimension may satisfy criteria on multiple axes (axis-inclusive sum) and is also reported deduplicated (unique). The primary encoder (pattern-deviation input, retinal coordinate-initialised positional encoding) allocates 50 of 128 distinct dimensions to spatial axes (70 axis assignments, since a dimension may satisfy criteria on more than one axis): 33 vertical midline, 18 horizontal raphe, and 19 central-peripheral. Per-encoder counts for the full 2×3 factorial are reported in eTable 1; the full per-dimension categorization for the primary encoder is provided in eTable 3.

**Supervised vertical-midline classifier — training, validation, and ablation.** A logistic regression classifier was fit on the 33 vertical-midline-categorized dimensions of the primary encoder, trained to separate expert-labeled neuro_suspect from not_neuro_suspect eye-fields in the UWHVF cohort. Validation used 5-fold patient-grouped stratified cross-validation. Hard-negative evaluation refers to the composition of the labeled set — all patients were identified by NHT screening — rather than to a per-field filter applied at validation. Held-out specificity was measured on two disjoint control sets of 50 patients each: clean normals (NHT < 10, MD > −3 dB) and clean glaucoma (NHT < 10, MD < −5 dB, low hemianopic loading on archetypal-pattern decomposition); both sets were excluded from classifier training. The top-20 operating threshold (signed log-odds ≥ 6.807) was set as the score cut-off corresponding to the 20th-ranked Harvard-GF patient under external application to Harvard-GF.

**Cross-modal VF-to-OCT decoder.** A two-layer multilayer perceptron (128 → 256 → 256; one 256-unit hidden layer, GELU activation; 98,816 parameters) was trained to predict 256-point peripapillary RNFL thickness profiles (TSNIT) from the primary encoder’s 128-dimensional VF embeddings. Training used Harvard-GF glaucoma-labeled patients with classifier rank below the top-200 threshold, with patient-level train/validation splits and the top-20 suspect cohort held out entirely. The mean-absolute-error prediction ceiling of approximately 23 µm clustered tightly across all 24 encoder × decoder combinations (six encoders × four decoder architectures), representing a representation-invariant ceiling reflecting the irreducible information gap between 52 VF locations and 256 peripapillary OCT positions under standard test–retest variability. Sectoral effect-size escape from the bootstrap null was computed using the same protocol as eTable 2.

**NHT empirical calibration.** The Boland NHT requires pattern deviation probability thresholds, which are proprietary to the Humphrey Field Analyzer. We estimated PD probability distributions empirically from 4,532 UWHVF normals (MD > −2 dB) at each of the 52 test locations. These empirical thresholds were used for both the UWHVF evaluation and Harvard-GF screening. We used the published Boland cutoff of 30 throughout.^17^

**Model architecture.** The encoder is a transformer with 4 layers, model dimension 128, 4 attention heads, feed-forward dimension 256, GELU activation, pre-norm LayerNorm, and dropout 0.1, initialized with Xavier uniform weights. Each of the 52 Humphrey 24-2 test locations is treated as a single token; no patch grouping is used. Input channels are projected to the model dimension by a linear layer before the first transformer block (1 channel for total-deviation or pattern-deviation encoders, 2 for the two-channel TD+PD encoders), so the two channels are fused before any attention operation. No classification token is used: the 128-dimensional representation is the mean of the 52 token outputs after the final encoder normalization. The reconstruction decoder is a 2-layer transformer of the same width, used only during pretraining and discarded thereafter. The encoder contains 537,088 parameters; encoder and decoder together contain 802,561.

**Positional encodings.** Positional encodings are learned parameters, initialized either to the retinal coordinates of each test location (primary encoder and the coordinate arm of the factorial) or randomly (random arm). A no-positional control was trained with positional encodings omitted entirely, to confirm that positional information is required; this control collapsed to severity-only features and reconstructed less accurately (mean absolute error 3.12 dB versus 1.96 dB for the primary encoder).

**Masking and pretraining objective.** During pretraining, a proportion of test locations sampled uniformly between 30% and 50% was masked per field; masking is applied to individual locations rather than contiguous blocks. Masked positions are zeroed at the encoder input and replaced by a learned mask token in the decoder. The training objective is a Huber loss (δ = 1.0) computed on masked locations only. For two-channel encoders the loss is averaged across the two output channels. Reconstruction performance is reported at a fixed mask ratio of 0.4.

**Training configuration and data splits.** Optimization used AdamW with learning rate 0.001, weight decay 0.05, a 20-epoch linear warmup followed by cosine decay to 0.00001, and batch size 256, for a maximum of 500 epochs with early stopping (patience 100) on validation reconstruction loss. The primary encoder stopped at epoch 339, with the selected checkpoint at epoch 239. Values were z-scored per location using training-split statistics, and Gaussian noise (σ = 0.2) was added to training inputs only. UWHVF was split by patient into 23,223 training, 2,866 validation, and 2,854 test fields, with a leakage assertion confirming no patient appears in more than one split. Training ran on Apple MPS hardware and took approximately 215 minutes. Analyses used Python with PyTorch, scikit-learn, and SciPy. No random seed was fixed in the training path, so reported results derive from a single training run and checkpoints are not bit-reproducible across runs.

**Cross-validation and the hard-negative design.** Cross-validation used 5-fold stratified splits grouped by patient identifier, so that all fields from a patient — both eyes and all longitudinal visits — fall within a single fold. The labeled evaluation set comprises 1,855 eye-fields from 231 patients (889 neuro_suspect from 221 eyes of 130 patients; 966 not_neuro_suspect from 184 eyes of 114 patients), with 13 patients contributing monocular-asymmetric eyes to both classes. Hard-negative evaluation refers to the composition of the labeled set rather than to a per-field filter: all patients in the set were identified by NHT screening (maximum NHT ≥ 30 on at least one field), so the negative class comprises eyes that triggered rule-based review but showed no neurological morphology on per-eye expert inspection. Individual fields within these patients span the full NHT range.

**Pathway-localized phenotype read-outs.** The ten pathway-localized phenotype read-outs shown in Figure 3 are hand-specified rather than fitted. For each phenotype, a set of latent dimensions was curated by inspection of dimension activation maps, and the score is the unweighted sum of those dimensions after z-scoring across the cohort; all weights are +1. No labels, model fitting, or cross-validation were used for these ten read-outs, and no discrimination performance is claimed for them. Cohorts were formed by ranking 13,052 bilateral UWHVF pairs by each score, assigning each eye to its highest-scoring phenotype, and taking the top 20 per phenotype; figure panels show the mean of the top 10. UMAP projections were computed on all 128 dimensions (n_neighbors = 30, min_dist = 0.1, euclidean metric, random_state = 17), fit on a 5,000-eye background sample with phenotype cohorts projected into that embedding. The anatomical pathway locations labeled in Figure 3 are clinical annotations of each phenotype's expected lesion site and are not outputs of the model.

**Feature-subspace and baseline comparisons.** To test whether classification performance depended on the anatomically interpretable vertical-midline dimensions rather than on feature count or access to the raw field, the classifier was compared, on identical folds and with the same logistic-regression head, against the complementary 95-dimension subspace, random 33-dimension subsets of the latent space, the raw 52-point pattern-deviation values, a principal-component decomposition of those values at full rank (52 components; a linear map to higher dimension cannot add information), and the NHT score alone. The vertical-midline subspace was selected for interpretability rather than unique sufficiency, and the primary encoder was selected on anatomical specialization rather than downstream accuracy.

### eFigures

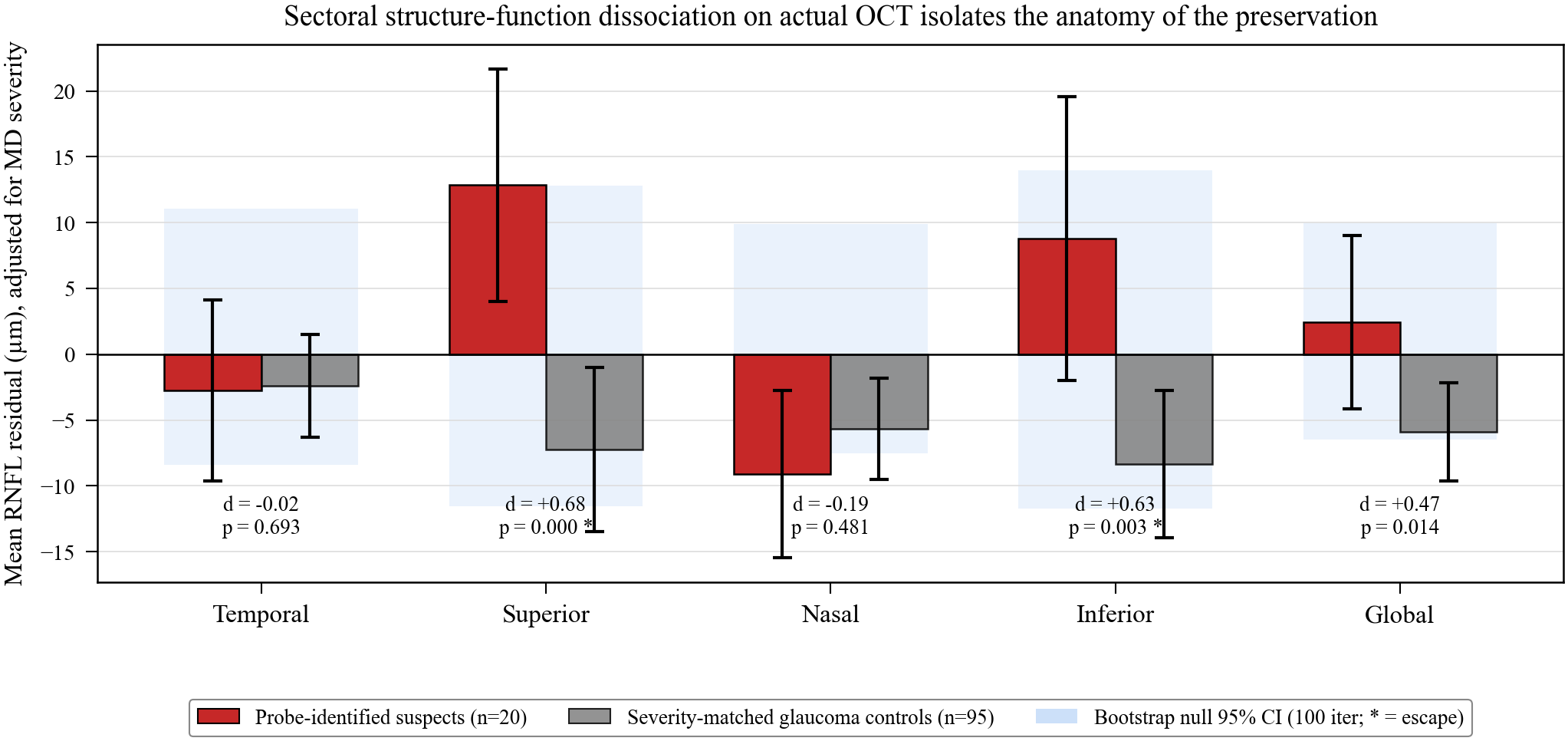

**eFigure 1:** Sectoral structure-function dissociation on actual OCT isolates the anatomy of the preservation. Sectoral RNFL residual (actual minus severity-adjusted prediction, µm) for classifier-identified top-20 suspects (n = 20) versus 95 severity-matched glaucoma controls, shown per peripapillary sector (temporal, superior, nasal, inferior) and globally. Bars show group means with 95% confidence interval; shaded horizontal bands show the 95% CI of a 100-iteration bootstrap null from random 20-patient MD-matched glaucoma subsets. Superior and inferior effects escape the null (Cohen's d = +0.68, p < 0.001 and d = +0.63, P = .003 respectively), while temporal and nasal sectors are null. The global effect, though nominally significant on Mann-Whitney (P = .014), does not escape the bootstrap null — the anatomy of the preservation (superior and inferior specifically) carries the discriminating signal rather than global RNFL magnitude. See eTable 2.

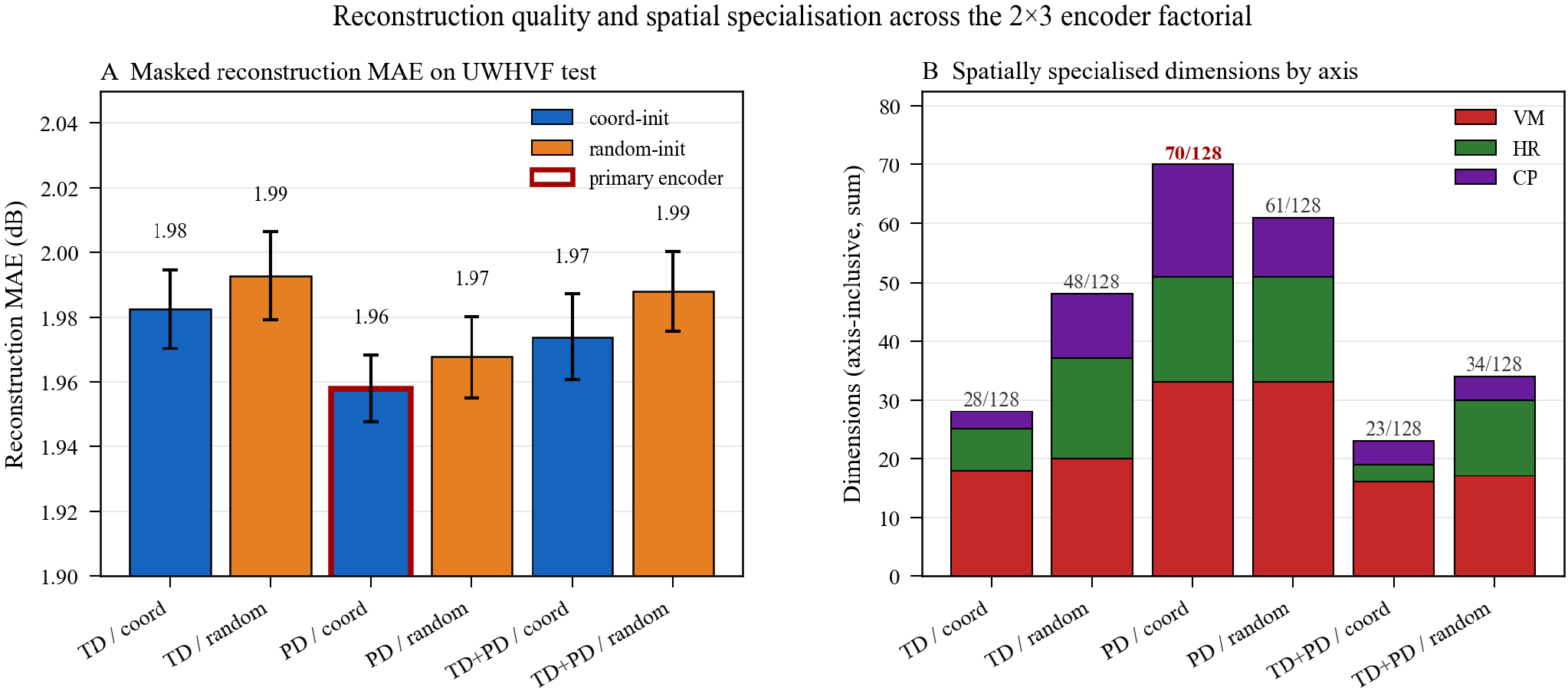

**eFigure 2.** Reconstruction quality and spatial specialization across the 2×3 encoder factorial. Panel A: masked-reconstruction MAE (mask ratio 0.4, 10-seed average) across all six encoders (TD / PD / TDPD × coordinate / random). Panel B: stacked per-axis spatially specialized dimension counts (axis-inclusive sum) across the same six encoders.

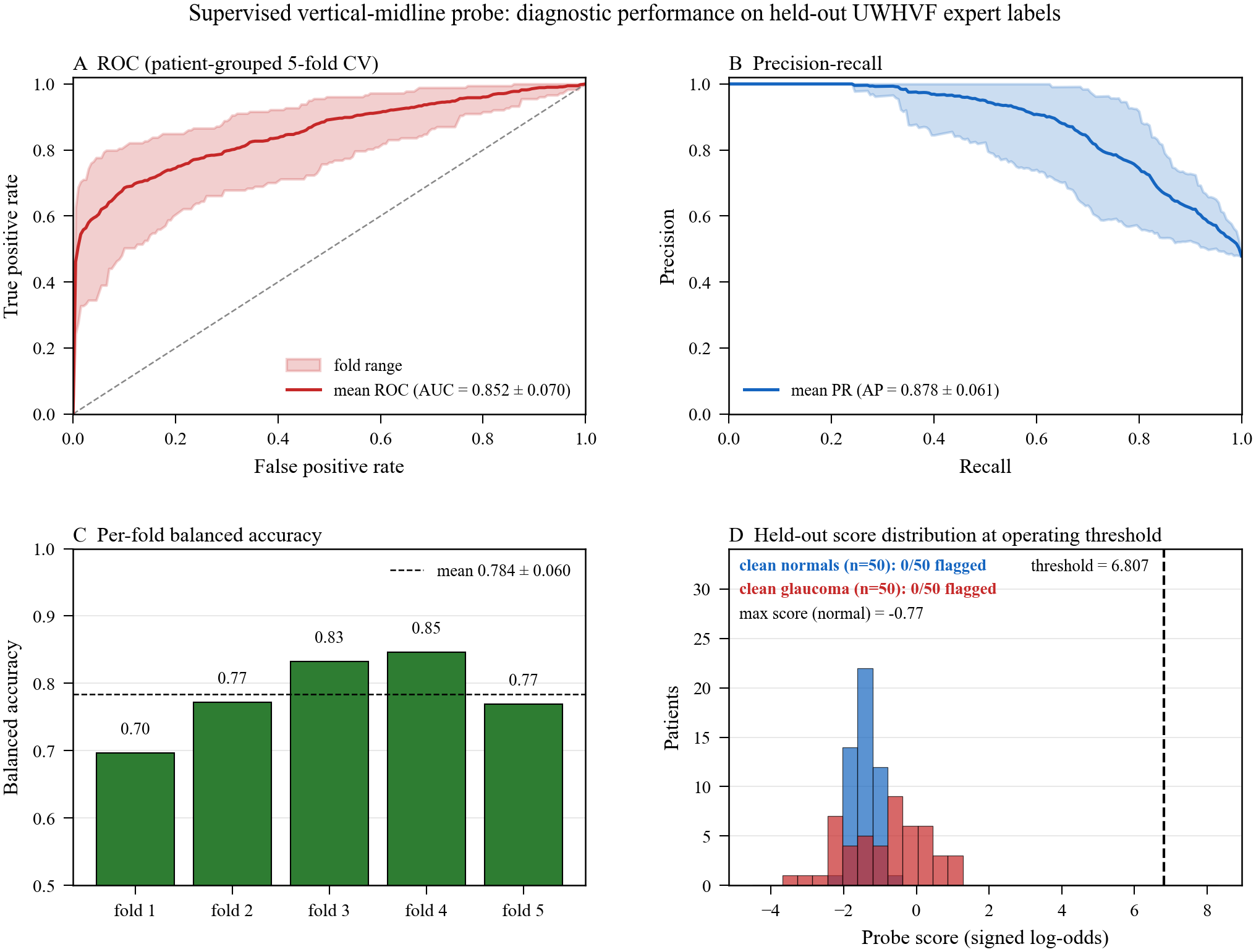

**eFigure 3.** Supervised vertical-midline classifier: diagnostic performance on held-out UWHVF expert labels. ROC, precision-recall, per-fold cross-validated balanced accuracy, and held-out specificity distribution on 50 clean normals and 50 clean glaucoma controls.

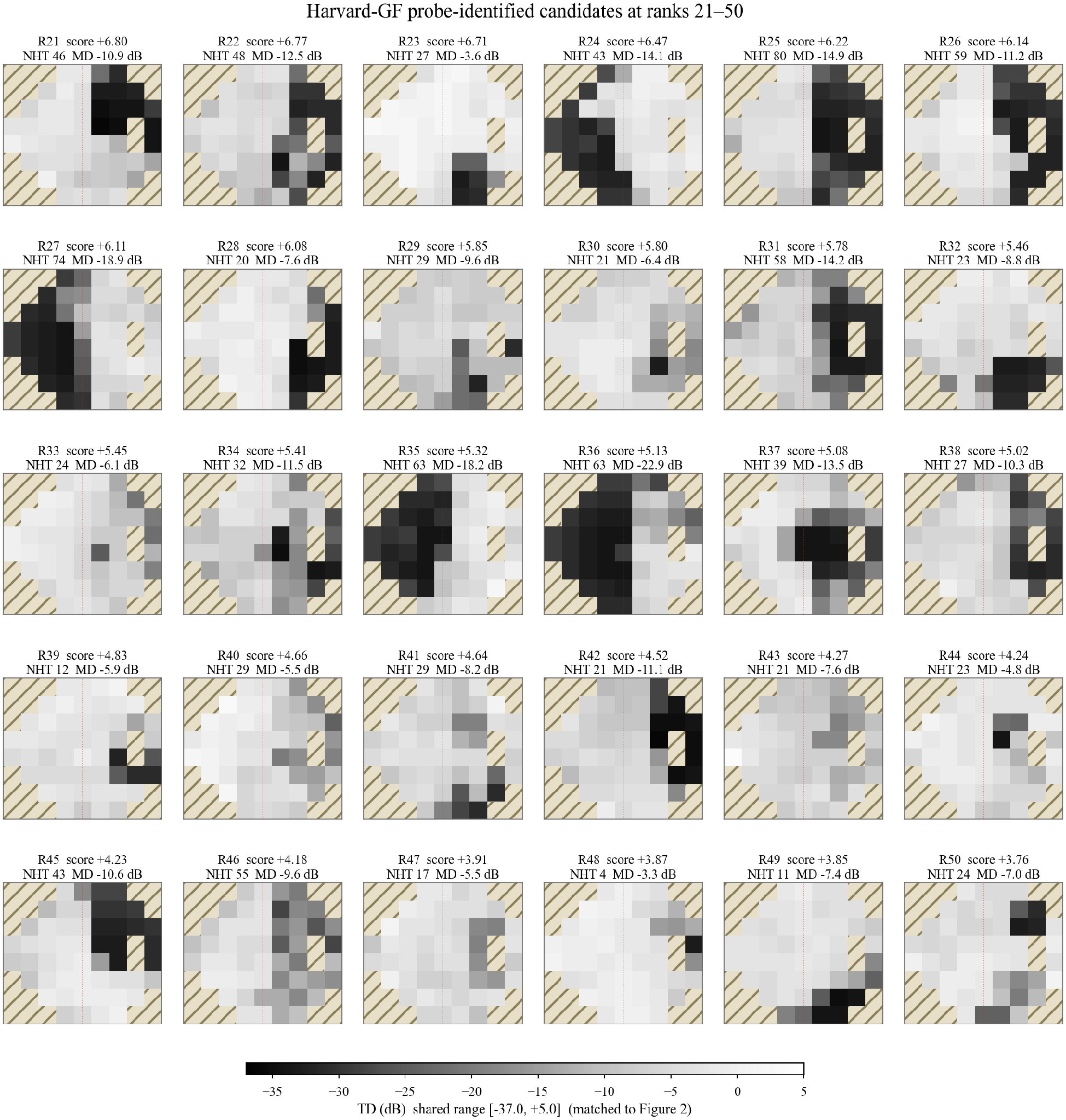

**eFigure 4.** Harvard-GF classifier-identified candidates at ranks 21–50. Total-deviation heatmap grid with per-patient rank, classifier score, NHT, and MD annotations.

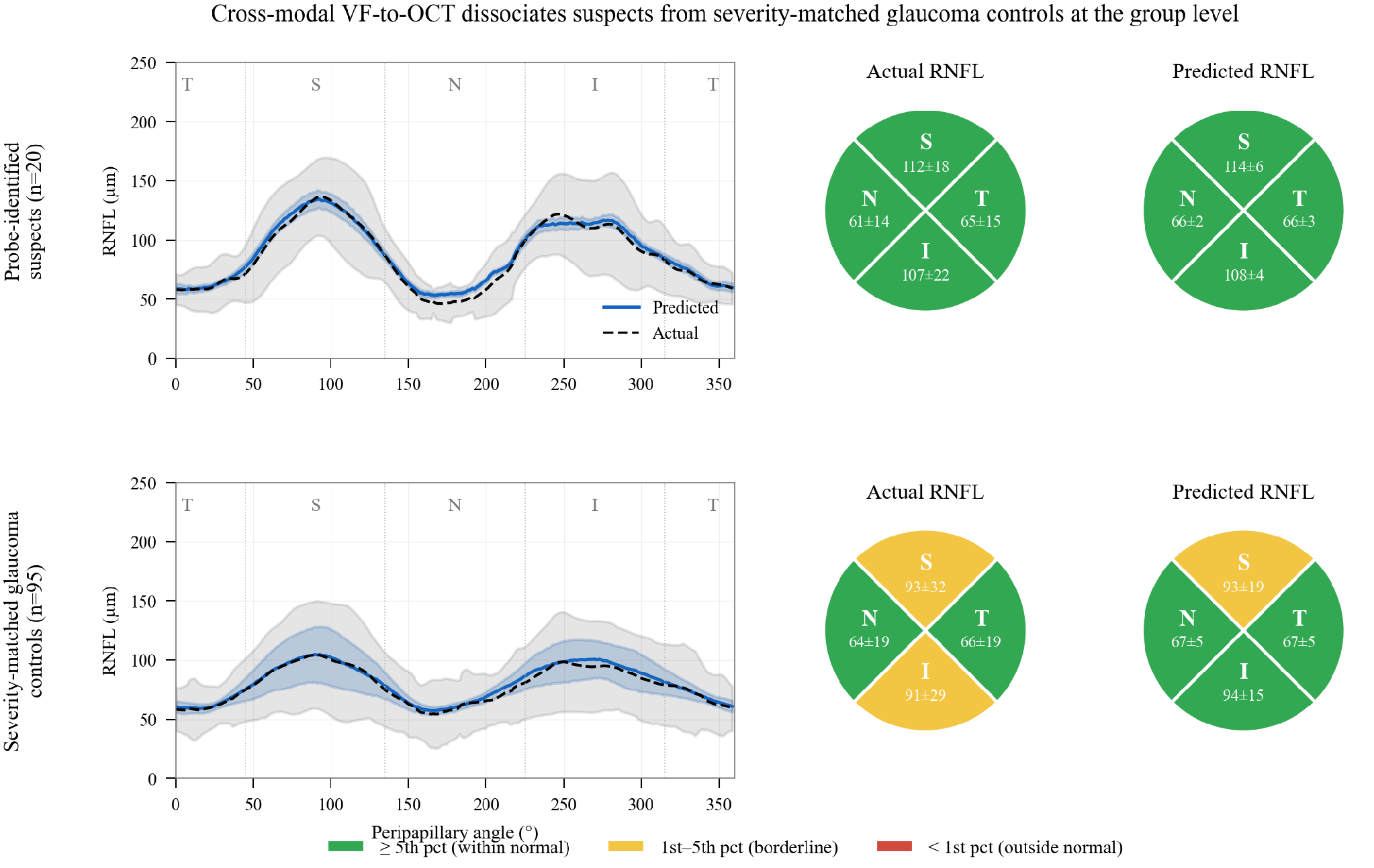

**eFigure 5.** Cross-modal VF-to-OCT reconstruction at the group level; the decoder was trained with all classifier-rank ≤ 200 patients excluded from training and validation. Top row: classifier-identified suspects (n=20); bottom row: severity-matched glaucoma controls (n=95). Each row shows the mean predicted (solid) and mean actual (dashed) peripapillary RNFL thickness as a function of angular position around the optic disc, with shaded ±1 SD bands; alongside, sector readouts of mean RNFL thickness as actual (left circle) and predicted (right circle) per-quadrant means, color-coded against Harvard-GF non-glaucoma reference percentiles (green ≥ p5, yellow p1–p5, red < p1). Suspect predicted and actual readouts converge on a preserved-RNFL prototype across all four sectors; severity-matched glaucoma controls show borderline thinning at the superior and inferior sectors on actual OCT, with the borderline superior sector reproduced in the predicted readout.

### eTables

#### eTable 1. Positional encoding ablation

|  | **TD, coordinate PE** | **PD, coordinate PE (primary)** | **TD+PD, coordinate PE** | **TD, random PE** | **PD, random PE** | **TD+PD, random PE** |
| --- | --- | --- | --- | --- | --- | --- |
| **Reconstruction MAE (dB)** | 1.98 | 1.96 | 1.97 | 1.99 | 1.97 | 1.99 |
| **Spatially specialized dimensions (axis-inclusive sum)** | 28 | 70 | 23 | 48 | 61 | 34 |
| of which: vertical midline | 18 | 33 | 16 | 20 | 33 | 17 |
| of which: horizontal raphe | 7 | 18 | 3 | 17 | 18 | 13 |
| of which: central–peripheral | 3 | 19 | 4 | 11 | 10 | 4 |
| **Unique spatial dimensions (deduplicated)** | 23 | 50 | 21 | 33 | 47 | 28 |

**eTable1:** Reconstruction MAE is masked-location mean absolute error on the UWHVF held-out test set (n = 2,854 fields; mask ratio 0.4; mean of 10 mask seeds; ±SD 0.01 dB across seeds for all encoders; TDPD values averaged across the TD and PD output channels). Dimension categorization tests each of 128 latent dimensions for differential activation between anatomically defined hemifield regions (nasal vs temporal for vertical midline; superior vs inferior for horizontal raphe; central vs peripheral for central–peripheral) at a 5 dB hemifield mean difference threshold; a single dimension may satisfy criteria on multiple axes. Axis-inclusive sum counts each qualifying dimension in every axis it satisfies (the count referenced in the main text). Unique dimensions deduplicates across axes. The six encoders constitute a 2×3 factorial design (input representation: TD / PD / TDPD × positional-encoding initialisation: retinal coordinate / random). The PD coordinate-initialised encoder is the primary encoder used for all downstream analyzes.

#### eTable 2. OCT structure-function preservation

| **Panel A — Actual OCT** | **Suspects (µm)** | **Controls (µm)** | **Δ µm (95% CI)** | **Cohen's d (95% CI)** | **p** | **Outside null 95% CI** |
| --- | --- | --- | --- | --- | --- | --- |
| **Temporal** | −2.8 ± 15.7 | −2.4 ± 19.3 | −0.4 (−8.5 to +6.9) | −0.02 (−0.44 to +0.42) | 0.69 | No |
| **Superior** | +12.9 ± 20.2 | −7.2 ± 30.9 | +20.1 (+9.3 to +30.8) | +0.68 (+0.31 to +1.15) | <0.001 | Yes |
| **Nasal** | −9.1 ± 14.5 | −5.7 ± 19.2 | −3.4 (−10.5 to +3.8) | −0.19 (−0.59 to +0.19) | 0.48 | No |
| **Inferior** | +8.8 ± 24.6 | −8.4 ± 27.9 | +17.2 (+4.9 to +28.5) | +0.63 (+0.15 to +1.14) | 0.003 | Yes |
| **Global** | +2.5 ± 15.0 | −5.9 ± 18.5 | +8.4 (+0.6 to +15.8) | +0.47 (+0.04 to +0.96) | 0.014 | No |
| **Panel B — Predicted TSNIT (cross-modal decoder)** |  |  |  | Predicted Cohen's d |  | Outside null 95% CI |
| **Temporal** |  |  |  | +1.59 |  | Yes |
| **Superior** |  |  |  | +1.40 |  | Yes |
| **Nasal** |  |  |  | +1.78 |  | Yes |
| **Inferior** |  |  |  | +1.37 |  | Yes |
| **Global** |  |  |  | +1.69 |  | Yes |

**eTable 2.** Panel A reports the OCT structure-function comparison between the classifier-identified top-20 Harvard-GF suspects and 95 unique severity-matched glaucoma controls (drawn from a 5:1 MD-matched pool of 100, deduplicated because MD-adjacent suspects share controls). Controls are defined as Harvard-GF glaucoma-labeled patients with classifier rank > 200 and NHT < 28, matched on MD (±2 dB). Sectors follow the Garway-Heath convention, defined by ±45° arcs around the cardinal peripapillary positions. RNFL residual = actual minus predicted from a glaucoma-specific linear regression of sectoral RNFL on MD, fit on the full Harvard-GF glaucoma cohort (n = 1,489 eyes). Δ and Cohen's d 95% confidence intervals are from 3,000-iteration non-parametric bootstraps; p values are Mann-Whitney U two-sided. Bootstrap-null test: a 100-iteration null distribution of Cohen's d was constructed by randomly sampling 20-patient pseudo-suspect subsets from the MD-matched glaucoma pool and computing the same sectoral comparison. Per-sector null 95% CIs were: temporal (−0.43, +0.57); superior (−0.37, +0.42); nasal (−0.39, +0.52); inferior (−0.42, +0.50); global (−0.35, +0.54). Observed Cohen's d in the superior and inferior sectors falls outside the null 95% CI (column 'Outside null 95% CI' = Yes). The global effect, though nominally significant on Mann-Whitney (P = .014) and with a d 95% CI excluding zero, does not fall outside the bootstrap null 95% CI and is therefore flagged No — the anatomy of the preservation (superior and inferior specifically) carries the discriminating signal, rather than global RNFL magnitude. Panel B reports the parallel sectoral analysis on RNFL profiles predicted from VF embeddings by the cross-modal decoder applied to the same 20-suspect cohort, with all patients of classifier rank ≤ 200 excluded from decoder training and validation. Predicted effects escape the bootstrap null in all four sectors and globally. Predicted preservation extends to the temporal and nasal sectors, where actual OCT shows no difference from controls (Panel A), indicating a morphology-level signal rather than sector-specific structural concordance.

#### eTable 3. Per-dimension axis categorization, primary encoder

| **Dimension** | **Vertical midline** | **Horizontal raphe** | **Central–peripheral** | **Category** |
| --- | --- | --- | --- | --- |
| 0 | Y | N | N | vertical-midline |
| 1 | Y | Y | N | multi-axis |
| 2 | N | N | N | severity |
| 3 | N | N | N | mixed |
| 4 | N | N | Y | central-peripheral |
| 5 | N | N | N | severity |
| 6 | N | N | N | severity |
| 7 | N | N | N | mixed |
| 8 | N | N | N | mixed |
| 9 | N | N | N | mixed |
| 10 | N | N | N | mixed |
| 11 | N | N | N | severity |
| 12 | N | N | N | mixed |
| 13 | N | Y | N | horizontal-raphe |
| 14 | N | Y | N | horizontal-raphe |
| 15 | N | N | N | severity |
| 16 | Y | N | Y | multi-axis |
| 17 | N | N | N | severity |
| 18 | Y | N | N | vertical-midline |
| 19 | Y | N | Y | multi-axis |
| 20 | Y | Y | Y | multi-axis |
| 21 | Y | N | N | vertical-midline |
| 22 | N | Y | Y | multi-axis |
| 23 | Y | Y | N | multi-axis |
| 24 | N | N | N | mixed |
| 25 | N | N | N | mixed |
| 26 | N | N | N | severity |
| 27 | N | N | N | severity |
| 28 | Y | Y | N | multi-axis |
| 29 | Y | N | N | vertical-midline |
| 30 | Y | N | N | vertical-midline |
| 31 | Y | N | N | vertical-midline |
| 32 | N | N | N | severity |
| 33 | N | N | N | mixed |
| 34 | N | N | N | severity |
| 35 | N | N | Y | central-peripheral |
| 36 | N | Y | Y | multi-axis |
| 37 | N | N | N | severity |
| 38 | N | Y | Y | multi-axis |
| 39 | Y | N | N | vertical-midline |
| 40 | N | N | N | mixed |
| 41 | Y | Y | N | multi-axis |
| 42 | N | Y | N | horizontal-raphe |
| 43 | N | N | N | severity |
| 44 | N | N | N | severity |
| 45 | Y | N | N | vertical-midline |
| 46 | N | N | Y | central-peripheral |
| 47 | Y | N | N | vertical-midline |
| 48 | Y | N | N | vertical-midline |
| 49 | N | Y | N | horizontal-raphe |
| 50 | N | N | N | severity |
| 51 | N | N | N | severity |
| 52 | N | N | N | severity |
| 53 | N | N | N | severity |
| 54 | N | N | N | severity |
| 55 | Y | N | N | vertical-midline |
| 56 | N | N | Y | central-peripheral |
| 57 | Y | N | Y | multi-axis |
| 58 | Y | N | N | vertical-midline |
| 59 | N | N | N | severity |
| 60 | N | N | N | severity |
| 61 | N | N | N | severity |
| 62 | N | N | N | severity |
| 63 | N | N | N | mixed |
| 64 | Y | N | N | vertical-midline |
| 65 | N | N | N | mixed |
| 66 | Y | Y | N | multi-axis |
| 67 | Y | Y | N | multi-axis |
| 68 | N | N | Y | central-peripheral |
| 69 | N | N | N | severity |
| 70 | Y | N | N | vertical-midline |
| 71 | N | N | N | mixed |
| 72 | N | N | N | severity |
| 73 | N | N | N | severity |
| 74 | N | N | N | severity |
| 75 | N | N | N | severity |
| 76 | N | N | N | mixed |
| 77 | N | N | N | mixed |
| 78 | N | N | N | mixed |
| 79 | N | N | N | severity |
| 80 | Y | N | N | vertical-midline |
| 81 | N | N | N | severity |
| 82 | N | N | N | mixed |
| 83 | N | N | N | severity |
| 84 | N | N | N | severity |
| 85 | N | N | N | severity |
| 86 | N | N | N | severity |
| 87 | N | N | N | severity |
| 88 | Y | N | N | vertical-midline |
| 89 | Y | N | Y | multi-axis |
| 90 | N | N | N | mixed |
| 91 | N | N | N | mixed |
| 92 | N | N | N | severity |
| 93 | N | N | N | mixed |
| 94 | Y | N | N | vertical-midline |
| 95 | N | N | N | severity |
| 96 | N | N | N | severity |
| 97 | N | N | Y | central-peripheral |
| 98 | N | Y | Y | multi-axis |
| 99 | N | N | N | severity |
| 100 | N | N | N | mixed |
| 101 | N | Y | N | horizontal-raphe |
| 102 | N | N | N | severity |
| 103 | N | N | N | mixed |
| 104 | N | N | N | severity |
| 105 | N | N | N | severity |
| 106 | N | N | Y | central-peripheral |
| 107 | N | N | N | mixed |
| 108 | N | N | N | mixed |
| 109 | N | N | N | severity |
| 110 | N | N | N | severity |
| 111 | N | N | N | severity |
| 112 | N | N | N | severity |
| 113 | N | N | N | severity |
| 114 | Y | N | N | vertical-midline |
| 115 | Y | Y | Y | multi-axis |
| 116 | N | N | N | severity |
| 117 | N | N | N | mixed |
| 118 | N | N | N | mixed |
| 119 | Y | N | Y | multi-axis |
| 120 | N | N | N | mixed |
| 121 | N | N | N | severity |
| 122 | N | N | Y | central-peripheral |
| 123 | N | N | N | mixed |
| 124 | N | N | N | severity |
| 125 | N | N | N | mixed |
| 126 | Y | Y | N | multi-axis |
| 127 | Y | N | N | vertical-midline |

**eTable 3.** Full per-dimension axis categorization for the primary encoder (pattern-deviation input, retinal-coordinate positional encodings). Each of the 128 latent dimensions was categorized by differential activation between anatomically defined hemifield regions; a dimension may satisfy criteria on more than one axis. Totals: 50 distinct spatially specialized dimensions; 70 axis assignments (33 vertical-midline, 18 horizontal-raphe, 19 central–peripheral); 78 dimensions without spatial specialization. Exclusive labels: vertical-midline only 19, horizontal-raphe only 5, central–peripheral only 8, multi-axis 18, severity 49, mixed 29. Severity denotes an absolute severity-activation difference greater than 10 dB without any spatial-axis flag; mixed denotes dimensions meeting no categorization threshold.

#### eTable 4. Expert-labeled evaluation set composition

**Overall composition**

| **Category** | **Fields** | **Eyes** | **Patients** |
| --- | --- | --- | --- |
| **neuro_suspect** | 889 | 221 | 130 |
| **not_neuro_suspect** | 966 | 184 | 114 |
| **uncertain (excluded from training)** | 785 | 157 | 100 |
| **Total reviewed** | 2,640 | 562 | 292 |
| **In cross-validation** | 1,855 | 405 | 231 |

**Per-fold composition (5-fold patient-grouped cross-validation)**

| **Fold** | **Patients** | **Fields** | **neuro_suspect fields** | **not_neuro_suspect fields** |
| --- | --- | --- | --- | --- |
| **Fold 1** | 46 | 368 | 177 | 191 |
| **Fold 2** | 46 | 368 | 177 | 191 |
| **Fold 3** | 46 | 369 | 176 | 193 |
| **Fold 4** | 45 | 374 | 179 | 195 |
| **Fold 5** | 48 | 376 | 180 | 196 |
| **Total** | **231** | **1,855** | **889** | **966** |

**eTable 4.** Composition of the expert-labeled UWHVF evaluation set used for classifier training and cross-validation. All patients were identified by NHT screening (score ≥ 30 on at least one field); every field from each flagged patient was reviewed per-eye. Thirteen patients contributed monocular-asymmetric eyes to both classes, so unique patient counts do not sum; eye and patient counts also overlap across categories because labels are assigned per eye-field. Cross-validation used 5-fold patient-grouped stratified splits (all fields from a patient in one fold). The 100 held-out specificity controls (50 clean-normal, 50 clean-glaucoma UWHVF patients) are disjoint from this set. Per-fold counts are reported in the lower panel.
