## Supplementary material for "CHIASM: A Self-Supervised Visual Field Encoder for Neuro-Ophthalmology": TRIPOD AI Checklist

Study type: development (D) and external evaluation (E) of a multivariable prediction model. Checklist per Collins GS, Moons KGM, Dhiman P, et al. BMJ 2024;385:e078378 (version 7 February 2024). Item wording is abridged from the published checklist. Locations refer to the main manuscript and Supplementary Materials; page numbers to be inserted from the final typeset PDF.

| **Section / Topic** | **Item** | **Checklist item** | **Reported on page** |
| --- | --- | --- | --- |
| **TITLE** | | | |
| Title | **1**  *D;E* | Identify the study as developing or evaluating the performance of a multivariable prediction model, the target population, and the outcome to be predicted | Title page (p 1); Abstract (p 2) |
| **ABSTRACT** | | | |
| Abstract | **2**  *D;E* | See TRIPOD+AI for Abstracts checklist | Abstract (pp 2–3) |
| **INTRODUCTION** | | | |
| Background | **3a**  *D;E* | Explain the healthcare context (including whether diagnostic or prognostic) and rationale for developing or evaluating the prediction model, including references to existing models | Introduction (pp 3–4) |
|  | **3b**  *D;E* | Describe the target population and the intended purpose of the prediction model in the context of the care pathway, including its intended users | Introduction (pp 3–4); Discussion (pp 11–12) |
|  | **3c**  *D;E* | Describe any known health inequalities between sociodemographic groups | Not reported. No demographic variables are model inputs; no sociodemographic inequality context described (see items 14, 23a) |
| Objectives | **4**  *D;E* | Specify the study objectives, including whether the study describes the development or validation of a prediction model (or both) | Abstract (p 2); Introduction, final paragraph (p 4); Methods (pp 5–6) |
| **METHODS** | | | |
| Data | **5a**  *D;E* | Describe the sources of data separately for the development and evaluation datasets, the rationale for using these data, and representativeness of the data | Methods, Datasets (pp 4–5); Limitations (p 12) |
|  | **5b**  *D;E* | Specify the dates of the collected participant data, including start and end of participant accrual; and, if applicable, end of follow-up | Not restated in manuscript; accrual periods are those of the source datasets, reported in refs 22 and 24 (Methods, Datasets, pp 4–5) |
| Participants | **6a**  *D;E* | Specify key elements of the study setting including the number and location of centres | Methods, Datasets (pp 4–5) |
|  | **6b**  *D;E* | Describe the eligibility criteria for study participants | Methods, Datasets and labelled-set paragraph (pp 4–5) |
|  | **6c**  *D;E* | Give details of any treatments received, and how they were handled during model development or evaluation, if relevant | Not applicable. Treatment data are not available in either public dataset and were not used |
| Data preparation | **7**  *D;E* | Describe any data pre-processing and quality checking, including whether this was similar across relevant sociodemographic groups | Methods (pp 5–6); eMethods (Suppl pp 1–3). Consistency across sociodemographic groups not assessed |
| Outcome | **8a**  *D;E* | Clearly define the outcome that is being predicted and the time horizon, including how and when assessed, the rationale for choosing this outcome, and whether assessment is consistent across sociodemographic groups | Methods (pp 5, 7); Limitations (p 12). Consistency across sociodemographic groups not assessed |
|  | **8b**  *D;E* | If outcome assessment requires subjective interpretation, describe the qualifications and demographic characteristics of the outcome assessors | Methods (pp 5, 7). Assessor demographic characteristics not reported |
|  | **8c**  *D;E* | Report any actions to blind assessment of the outcome to be predicted | Methods (p 7); eMethods (Suppl p 3) |
| Predictors | **9a**  *D* | Describe the choice of initial predictors and any pre-selection of predictors before model building | Methods (pp 6–7) |
|  | **9b**  *D;E* | Clearly define all predictors, including how and when they were measured (and any actions to blind assessment) | Methods (pp 5–6); eMethods (Suppl pp 1–2). Predictors are device-measured perimetric values obtained before and independent of labels |
|  | **9c**  *D;E* | If predictor measurement requires subjective interpretation, describe the qualifications and demographic characteristics of the predictor assessors | Not applicable. Predictors are instrument outputs; no subjective interpretation |
| Sample size | **10**  *D;E* | Explain how the study size was arrived at, and justify that it was sufficient. Include details of any sample size calculation | No formal sample size calculation; all available public data were used. Precision indicated by patient-level bootstrap CIs (Table 1, pp 13–14) and exact binomial bounds |
| Missing data | **11**  *D;E* | Describe how missing data were handled. Provide reasons for omitting any data | Both datasets distribute complete 52-location fields; complete-case by construction. Eyes labelled uncertain on expert review were excluded (eMethods, Suppl p 3; eTable 4, Suppl p 14) |
| Analytical methods | **12a**  *D* | Describe how the data were used in the analysis, including whether the data were partitioned, considering any sample size requirements | Methods (pp 5–6, 8); eMethods (Suppl pp 2–3) |
|  | **12b**  *D* | Describe how predictors were handled in the analyses (functional form, rescaling, transformation, or standardisation) | Methods (p 6); eMethods (Suppl p 2) |
|  | **12c**  *D* | Specify the type of model, rationale, all model-building steps including hyperparameter tuning, and method for internal validation | Methods (pp 6–7); eMethods (Suppl pp 2–3) |
|  | **12d**  *D;E* | Describe if and how heterogeneity in model parameter values and performance was handled and quantified across clusters | Not examined. Single-source data at each stage; no multi-centre pooling |
|  | **12e**  *D;E* | Specify all measures and plots used (and their rationale) to evaluate model performance and, if relevant, to compare multiple models | Methods, Statistical Methods (p 8); Table 1 (pp 13–14); eFigure 3 (Suppl p 6). Calibration not assessed: the model is applied as a fixed-threshold ranking and audit tool, not as a probability estimator |
|  | **12f**  *E* | Describe any model updating arising from the model evaluation | Not applicable. No updating or recalibration was performed |
|  | **12g**  *E* | For model evaluation, describe how the model predictions were calculated | Methods, Cross-dataset screening (p 7); Data Sharing Statement (p 24) |
| Class imbalance | **13**  *D;E* | If class imbalance methods were used, state why and how this was done, and any subsequent recalibration | Not applicable. No imbalance methods used; classes near-balanced (889 vs 966 fields); balanced accuracy reported |
| Fairness | **14**  *D;E* | Describe any approaches that were used to address model fairness and their rationale | Not performed. No demographic variables enter the model and no subgroup analyses were conducted |
| Model output | **15**  *D* | Specify the output of the prediction model. Provide details and rationale for any classification and how thresholds were identified | Methods (p 7); Table 1 (pp 13–14); Discussion (p 11) |
| Development vs evaluation | **16**  *D;E* | Identify any differences between the development and evaluation data in healthcare setting, eligibility criteria, outcome, and predictors | Methods, Datasets (pp 4–5); Discussion (p 11); Limitations (p 12) |
| Ethical approval | **17**  *D;E* | Name the institutional research board or ethics committee that approved the study and describe consent or waiver | Methods, Datasets, final sentence (p 5): analysis of publicly available, de-identified datasets; not human subjects research requiring IRB approval |
| **OPEN SCIENCE** | | | |
| Funding | **18a**  *D;E* | Give the source of funding and the role of the funders for the present study | Funding/Support; Role of the Funder/Sponsor (pp 23–24) |
| Conflicts of interest | **18b**  *D;E* | Declare any conflicts of interest and financial disclosures for all authors | Conflict of Interest Disclosures (p 23) |
| Protocol | **18c**  *D;E* | Indicate where the study protocol can be accessed or state that a protocol was not prepared | No study protocol was prepared |
| Registration | **18d**  *D;E* | Provide registration information for the study, or state that the study was not registered | The study was not registered |
| Data sharing | **18e**  *D;E* | Provide details of the availability of the study data | Data Sharing Statement (p 24) (UWHVF, BSD 3-Clause; Harvard-GF, CC BY-NC-ND 4.0; expert-labelled evaluation set from the corresponding author) |
| Code sharing | **18f**  *D;E* | Provide details of the availability of the analytical code | Data Sharing Statement (p 24) |
| **PATIENT & PUBLIC INVOLVEMENT** | | | |
| Patient & public involvement | **19**  *D;E* | Provide details of any patient and public involvement, or state no involvement | No patient or public involvement in the design, conduct, reporting, interpretation, or dissemination of this study |
| **RESULTS** | | | |
| Participants | **20a**  *D;E* | Describe the flow of participants through the study, including numbers with and without the outcome. A diagram may be helpful | Figure 1 (legend p 15) and Methods (pp 4–5, 7) |
|  | **20b**  *D;E* | Report characteristics overall and for each data source, including key dates, key predictors, sample size, number of outcome events, and missing data | Table 1 (pp 13–14); eTable 2 (Suppl p 8); eTable 4 (Suppl p 14). Severity and screening characteristics reported; demographic characteristic tables not reported (see item 14) |
|  | **20c**  *E* | For model evaluation, show a comparison with the development data of the distribution of important predictors | Methods, Datasets (p 5); Table 1 (pp 13–14). Full side-by-side distribution table not provided |
| Model development | **21**  *D;E* | Specify the number of participants and outcome events in each analysis | Methods (pp 4–8); Table 1 (pp 13–14); eMethods (Suppl pp 1–3); eTable 4 (Suppl p 14) |
| Model specification | **22**  *D* | Provide details of the full prediction model to allow predictions in new individuals and third-party evaluation, including any access restrictions | eMethods (Suppl pp 2–3); eTable 3 (Suppl pp 9–13); Data Sharing Statement (p 24) |
| Model performance | **23a**  *D;E* | Report model performance estimates with confidence intervals, including for any key subgroups | Table 1 (pp 13–14); Findings (pp 8–10); eFigure 3 (Suppl p 6). Sociodemographic subgroup performance not assessed (see item 14) |
|  | **23b**  *D;E* | If examined, report results of any heterogeneity in model performance across clusters | Not examined (see item 12d) |
| Model updating | **24**  *E* | Report the results from any model updating, including the updated model and subsequent performance | Not applicable. No updating was performed |
| **DISCUSSION** | | | |
| Interpretation | **25**  *D;E* | Give an overall interpretation of the main results, including issues of fairness, in the context of the objectives and previous studies | Discussion (pp 10–12). Fairness not discussed (see item 14) |
| Limitations | **26**  *D;E* | Discuss any limitations of the study and their effects on biases, statistical uncertainty, and generalisability | Limitations (p 12) |
| Usability in current care | **27a**  *D* | Describe how poor quality or unavailable input data should be assessed and handled when implementing the model | Not reported. Handling of unreliable fields at deployment is not addressed in this version |
|  | **27b**  *D* | Discuss whether users will be required to interact in the handling of the input data or use of the model, and what expertise is required | Discussion, virtual-clinic paragraph (p 11) |
|  | **27c**  *D;E* | Discuss any next steps for future research, with a specific view to applicability and generalizability of the model | Conclusions (p 13); Limitations (p 12) |
